# Variation in Evening Light Exposure Pattern Between Myopes and Non-Myopes

**DOI:** 10.64898/2026.01.25.26344801

**Authors:** Kavya P. Katragadda, Rohit Dhakal, Pavan K. Verkicharla

## Abstract

**Purpose:** Considering a broad range of ambient lighting conditions may impact myopia development and dim light exposure as a potential risk factor for myopia progression, this study investigates i) light exposure pattern of Indian schoolchildren in evening hours, and ii) difference in evening light exposure patterns between myopic and non-myopic schoolchildren.

**Method:** 136 Indian schoolchildren aged 9-15 years, with 46 myopes and 90 non-myopes were recruited. This study analyzed evening light exposure (6-10:00 pm) recorded using MyLyt wearable light trackers in 136 Indian schoolchildren (age range: 9-15 years). Statistical analyses were conducted to evaluate differences in average light exposure levels, maximum light exposure levels, and time spent at each various light exposure (≤30 lux, >30 lux, ≤100 lux, and >100 lux across all the tested days) in myopic (n=46) and non-myopic (n=90) groups.

**Results:** The median evening light exposure level across all participants was 27 lux, with participants spending only 2 [0-6] minutes (median [IQR]) in light levels >100 lux. Myopic children had significantly lower median light exposure levels than non-myopic children (24 [19-30] vs. 28 [21-42] lux, p=0.017), and significantly lower maximum evening light levels than non-myopes (162 [101-273] lux vs. 189 [129-396] lux, p=0.044). Additionally, myopes spent significantly less time in light levels >30 lux than non-myopes (18 [11-31] minutes vs. 30 [15-53] minutes, p=0.009).

**Conclusions:** Indian schoolchildren spend evening hours in dim light <30 lux, below recommended illuminance for reading or studying. The significant differences in evening light exposure between myopic and non-myopic children needs further exploration for its causal relationship with myopia.

## Introduction

With the rising prevalence and severity of myopia, it is important to evaluate potential modifiable factors for moderating myopia incidence and progression [1]. One such factor is outdoor light exposure, which has been shown to attenuate myopia in animal model studies and is negatively associated with severity of myopia in epidemiologic studies [2, 3, 4-7]. Although myopia is considered to be pandemic, geographical and cultural diversities have likely to have attributed to a heterogenous distribution of myopia prevalence across the world. Read et. al found that Australian children had significantly longer outdoor light exposure than Singaporean children; this outcome within the context of Singapore’s relatively high myopia rates and Australia’s comparatively low myopia rates worldwide suggests that differences in children’s lifestyles governing outdoor light exposure may play a role in the country specific myopia prevalence [5]. In another study, Read et. al used wearable wrist mounted light trackers to monitor light exposure levels in a cohort of Australian children aged 10-15 years and reported that myopic children spent significantly less time (p < 0.001) at outdoors in light levels >1000 lux than their non-myopic peers [6]. In contrast, another study comparing light exposure levels in Chinese myopic and non-myopic children found similar average daily light exposure duration (p = 0.13) under light intensities of >1000 lux [7].

While protective effect of high-intensity daylight exposure against myopia, there is less evidence surrounding the range of dim light exposure levels as risk or protective factors against myopia development or progression. Experiments conducted in animal models have demonstrated that functional rod photoreceptors are essential to develop experimentally induced myopia [2]. Interestingly, experiments showed that both the photopic and scotopic light levels prevented lens induced myopia with increases in dopamine turnover, whereas mesopic light levels (50 lux) increased the experimental myopia with a decrease in dopamine turnover. Additionally, a study by Landis et. al in a cohort of Australian children found that increased time spent in mesopic light was correlated with more severe myopia [8]. Dhakal et al. investigated light exposure patterns of Indian schoolchildren aged 9-15 years during daytime (7:30 to 18:00 clock hours) and reported significant difference in illuminance exposure, time spent outdoors and epoch between myopes and non-myopes during transition or break time during day [4].

Considering that a broad range of ambient lighting conditions in entire day of human may potentially impact development of myopia, and that understanding light exposure patterns outside school hours when children have more control of their activities is important, this study aimed to investigate i) light exposure pattern of schoolchildren in evening hours in Indian children, and ii) if there is any significant difference in evening light exposure patterns between myopic and non-myopic schoolchildren. We hypothesize that myopic schoolchildren would spend comparatively more time in dim light than non-myopic children, a difference that could both contribute to and exacerbate myopia.

## Methods

This is an analysis part of a larger dataset collected as part of “Light Outdoor Myopia Study (LOMS)” conducted in two schools of Hyderabad, India, which had similar daily school schedules [4]. The study focused on children aged 9-15 years and reported baseline findings from the data collected in November and December 2021. The study adhered to the tenets of the Declaration of Helsinki and was approved by the Institutional Review Board of L V Prasad Eye Institute, Hyderabad, India (LEC 10-19-354). Written informed consent was obtained from the school principal and parents of each participant (children).

The previous study included 143 schoolchildren where unaided monocular distance visual acuity was measured using a logMAR chart, and refractive error was determined with an open-field auto-refractor (Shin Nippon, NVision-K 5001, Japan). Participants were then categorized as myopes or non-myopes based on their visual acuity and spherical equivalent refraction (SER). Participants qualified as myopes if their unaided visual acuity was worse than or equal to 0.1 logMAR and SER of ≤ −0.75 D with the sphere being ≤ −0.50 D, in either or both eyes. Participants from both the myopic and non-myopic groups were given personalized wearable “MyLyt” trackers to record real-time light exposure levels (lux). In short, a chest-mounted validated MyLyt tracker was used to objectively quantify light exposure profiles of schoolchildren. The tracker was clipped onto the school uniform (during school time) or the casual wear (outside of school time) worn by the participant, below the neck at the level of upper thoracic region. They were instructed to immediately relocate trackers onto new clothes after changing the school uniform. Further methods details are provided in our previous publication (see Dhakal et al., [4]). The MyLyt tracker has the capacity to record data continuously for a maximum of 7 days at a sampling rate of 1 data/min when fully charged. To minimize loss of data collection, a criterion of six consecutive days was used. Evenings were counted when participants had light exposure data > 30 minutes of consecutive data. Each included participant had a minimum of 1 evening with consecutive data and a maximum of 6 evenings. 79% of participants had 3 or greater evenings of data, and myopic participants had an average of 4.02 days with available data while non-myopic participants averaged 3.6 days. For this study, the data analyses include assessment of light exposure levels from 6:00 pm to 10:00 pm in the evening, which is after the average sunset time in the months of November and December in Hyderabad. A total of 143 participants consisting of 47 myopes and 96 non-myopes made up the original study subject group. Seven participants were excluded due to inappropriate data recording in the evening, such that a total of 136 participants data were analyzed in this study. Both school day and non-school day data were combined for the analysis.

### Statistical Analysis

Data previously logged by the MyLyt tracker were exported into Microsoft Excel 2023 (Microsoft Corporation, USA), where data was filtered values between (6-10:00 pm data only) and cleaning was done. Statistical analyses were performed using Excel and IBM SPSS Statistics 29.0.2.0 (IBM Corporation, USA). The average and the maximum evening lux levels were identified for each participant. Likewise, the time spent at various light levels ≤30 lux (mesopic), >30 lux (indoor photopic), ≤100 lux, and >100 lux was counted and was considered for all the days for each participant. The cutoff of 30 lux was chosen based on the median illuminance exposure level across participants and also because light levels <30 lux correspond to mesopic light levels based on previously-established guidelines [8]. For each participant, the average light exposure level was calculated by adding all the available light exposure level data in lux from 6-10:00 pm and dividing by the total number of evening lux levels logged across all available days. Maximum light exposure level was calculated across all days in which participants had data from 6-10:00 pm. Time spent in different light level categories was calculated by adding all lux values within the light level category (i.e., for the category of lux ≤30, all values ≤30 lux were added across all evenings with available data), yielding total minutes spent at a given light level category. Given the variability in the number of days that each participant had data for (depending on tracker wearing habits and the varied timeframes in which trackers were distributed to different cohorts), the average minutes spent at each light level category per evening was calculated. The total minutes each participant spent at each light level category across all days was divided by the number of days with data for each participant, resulting in an average time spent (in minutes) at each light level category. This process was repeated for lux levels of ≤30, >30, ≤100, and >100 lux. While 100% of the 136 participants had data for time spent in >30 lux and 80% of participants spent time in 100-199 lux, there was a steep drop off in light tracker data for light levels > 200 lux, with just 42% of participants having available data. As such, these categories were excluded from the final analysis. A sub-analysis comparing average light exposure levels, and time spent at different light levels were conducted between myopic and non-myopic participants.

Normality tests including Shapiro-Wilk tests, skewness and kurtosis determined that all parameters: average light exposure level, maximum light exposure level and time spent at different light exposure levels were not normally distributed. The data were compared between myopic (46) and non-myopic (90) subgroups using Mann-Whitney U tests. A p value of <0.05 was defined as significant, and data are represented as median [IQR: Q1-Q3].

## Results

### Illuminance exposure level

Across 136 participants, the median [IQR] illuminance exposure level between 6-10:00 pm was 27 [21-38] lux with a significant difference in median light exposure levels (p=0.017) between myopes (24 [19-30] lux) and non-myopes (28 [21-42] lux) as shown in Fig 1. The maximum median illuminance exposure level across all 136 participants was 183 [117-330] lux with the lowest value being 17 lux and the highest being 1,975 lux (Fig 2). There was also a significant difference (p=0.04) in maximum light exposure levels between myopic and non-myopic participants (162 [101-273] lux for myopes and 189 [129-396] lux for non-myopes).

**Fig 1.**
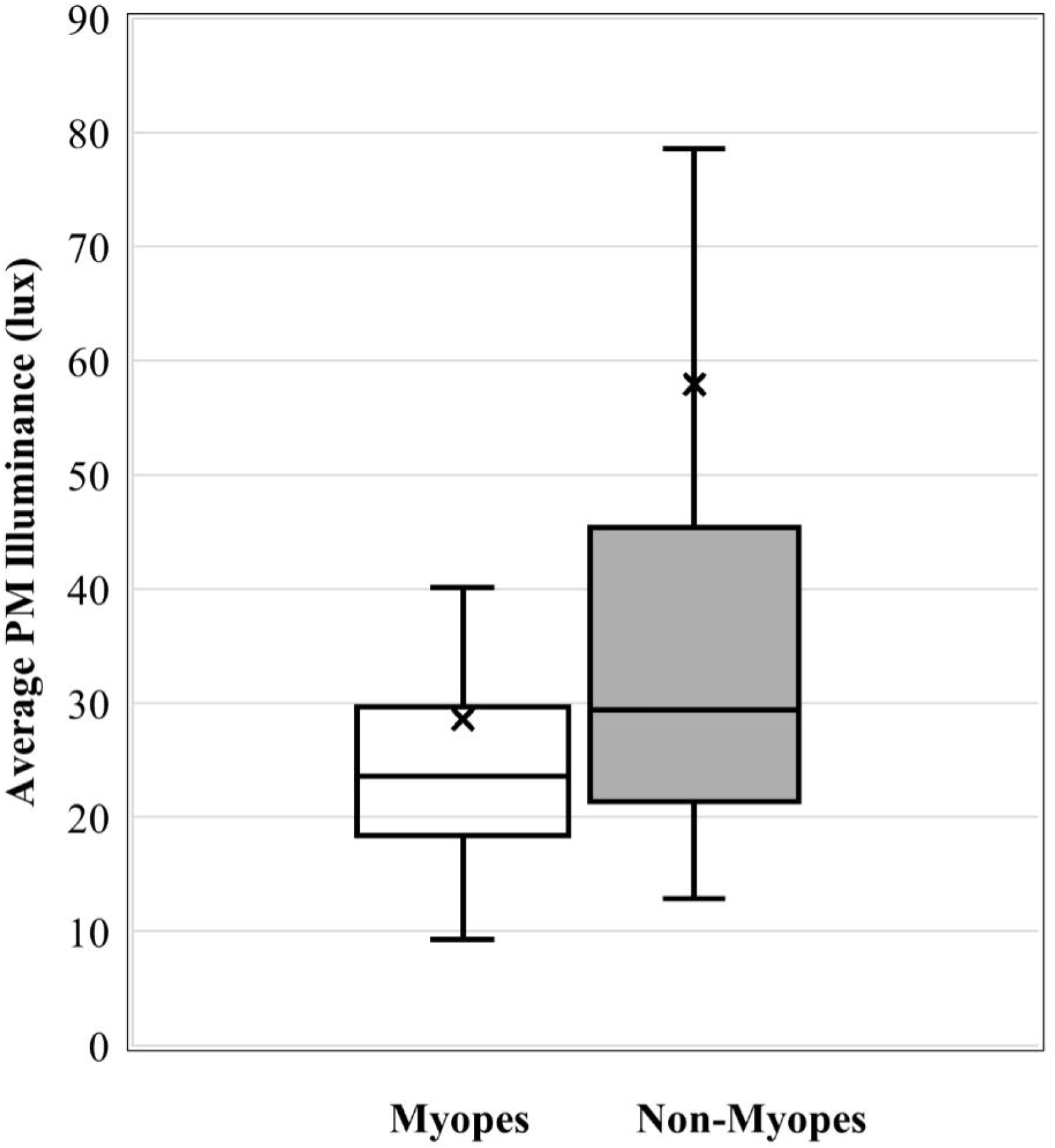
Distribution of evening (PM) average illuminance exposure level between myopic and non-myopic schoolchildren. X represents the average of the distribution. * Denotes statistical significance of <0.05 between data.

**Fig 2.**
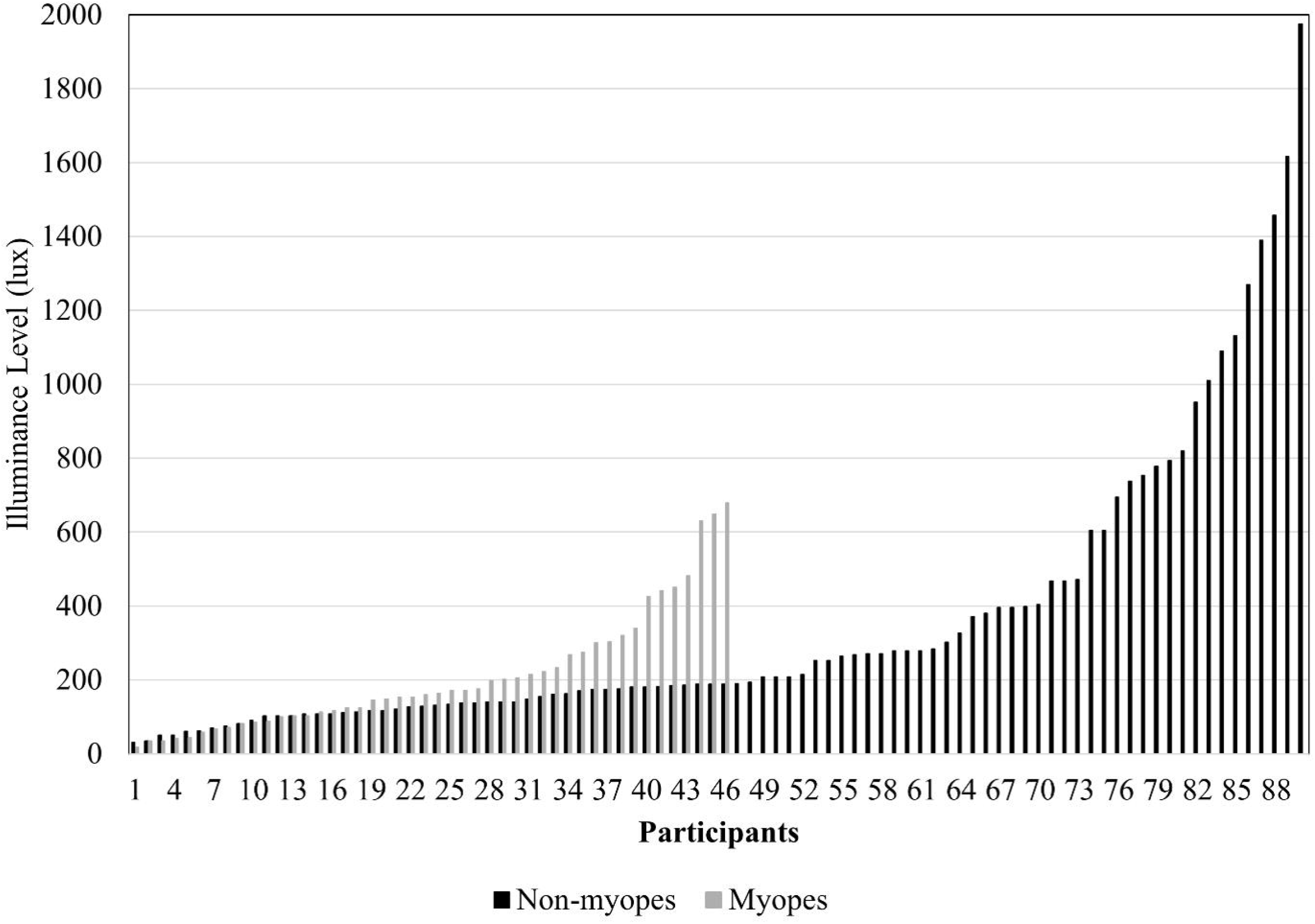
Distribution of maximum illuminance exposure (lux) level recorded in myopic and non-myopic participants. Note that we had 46 myopes and 90 non-myopes, and the data is arranged in ascending order to show the trend.

### Time Spent at Different Light Levels

From 6-10:00 pm, participants spent 75 [58-97] minutes per evening in light levels less ≤30 lux, 24 [12-43] minutes per evening in light levels > 30 lux, and just 2 [0-6] minutes in light levels >100 lux. There were small but significant differences in light exposure levels between myopes and non-myopes (Fig 3). Myopes spent lesser time in light levels >30 lux than non-myopes (18 [11-31] minutes versus 30 [15-53] minutes, p=0.009). A similar trend was noted for lux levels >100 lux (1 [0.1-4] minutes versus non-myopes (3 [1-7] minutes, p=0.013). There was no significant difference in time spent in light levels ≤30 lux and time spent in light levels ≤100 lux between myopes and non-myopes.

**Fig 3.**
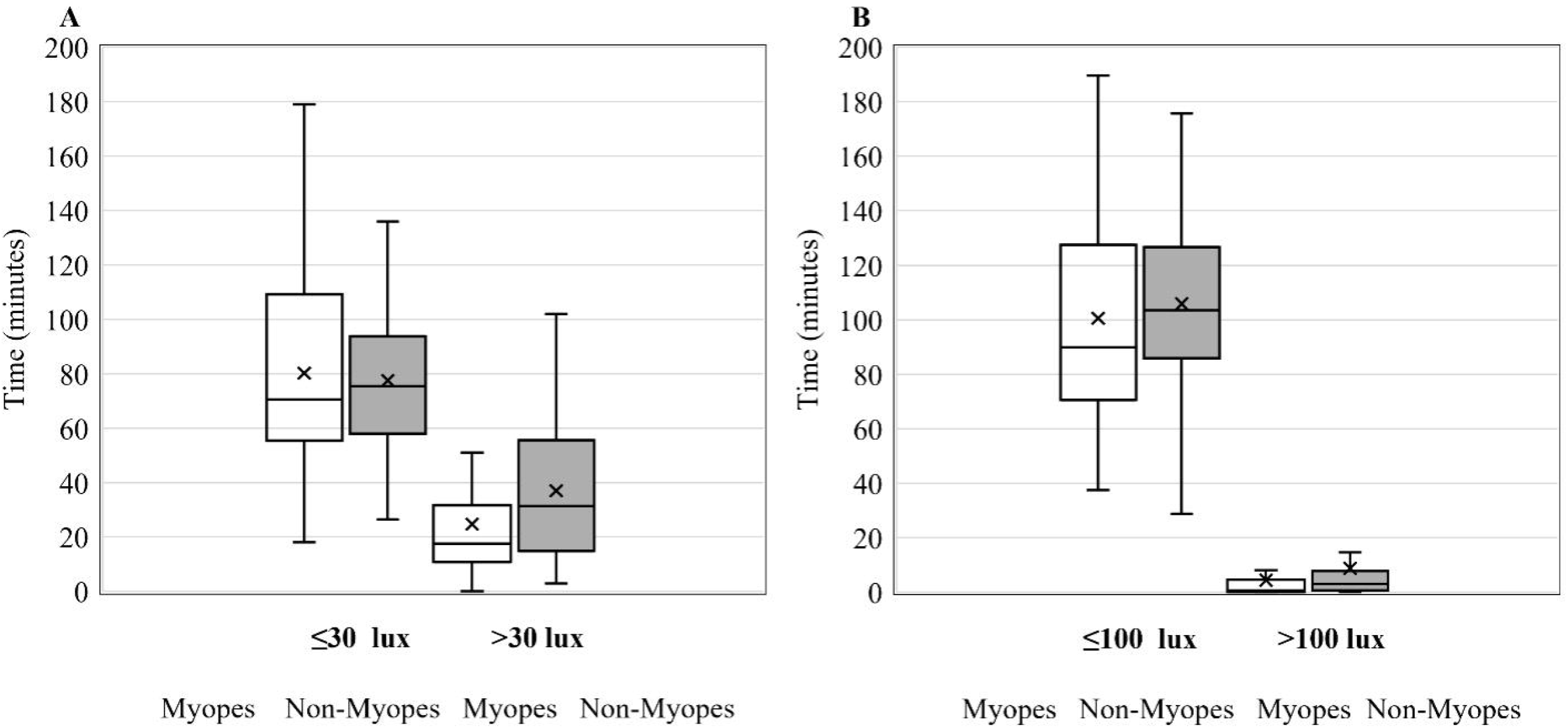
Distribution of time spent at different light levels by myopes and non-myopes in the evening. Panel A shows time spent across ≤30 and >30 lux, whereas panel B shows time spent across ≤100 and >100 lux. X represents the average of the distribution * represents statistical significance of <0.05 between data.

## Discussion

This study is the first to evaluate the evening light exposure levels in Indian schoolchildren. Key results from this study showed that Indian schoolchildren are exposed to a low light level and spend minimal time in light levels >100 lux in evenings. Between 6-10:00 pm, participants had median of just 2 minutes in light levels >100 lux and were exposed to a median illuminance exposure level of 27 lux, with a median maximum evening light exposure of <200 lux across all participants. Children with myopia overall had significantly lower evening light exposure levels, spent significantly less evening time than in light levels >30 lux and >100 lux than non-myopic children.

A study by Landis et. al on dim light exposure in Australian children examined the patterns of scotopic (<1-1 lux), mesopic (1-30 lux), indoor photopic (>30-1,000 lux) and outdoor photopic light (>1,000 lux) exposure in myopic and non-myopic participants, during the day [8]. Our study conducted to investigate light exposure levels in evening/ after school hours reveals that myopes spend more time in dim light than non-myopes, with significantly lower median (p=0.017) and maximum (p= 0.044) light exposure in lux between 6-10:00 pm. Non-myopes spent 1.7 times as many minutes in photopic lighting >30 lux and 4.2 times as many minutes in photopic lighting >100 lux as myopes, which is a small but significant difference (P=0.009, P =.013). These results suggest a correlation between mesopic lighting exposure/time spent in mesopic lighting and refractive error status in Indian schoolchildren, which is consistent with a finding from Landis et. al that myopic Australian children spend more time in mesopic light [8]. This concordance could suggest further evidence for the association between mesopic light exposure levels and myopia status, despite the potential differences in activity and behavioral patterns of populations across different nations.

This study examined dim light exposure levels in evenings alone rather than analyzing dim light exposure levels over an entire day. Importantly, the finding reveal that Indian schoolchildren spend much of their evenings in poorly lit, mesopic lighting conditions < 30 lux. The outcomes are important because mesopic lighting has been found to shift refractive error towards myopia and in animal experimental subjects, has been correlated with an exaggeration of axial growth, suggesting exacerbation of myopia progression and development [2, 9]. Furthermore, considering Indian children attend school until 4-5 pm for 6 days weekly and often spend evenings doing homework or in tuition, children should be in environments with light levels that are, at a minimum, 6x the median light exposure in lux found in this study. The recommended lighting for bedrooms is 50-150 lux, for casual reading is 150 lux, and for studying is 150-750 lux – much higher than the median evening light level exposure of 27 lux reported from this study [12].

This low evening light exposure level exposure may provide support for the theory that Indian subjects may be exposed to lower light levels in general, compared to reported data from other countries. Dhakal et. al found that myopes were exposed to an average of 789 [511] lux/day, while non-myopes were exposed to an average of 908 [466] lux/day [4]. This level of light exposure is lower than average light levels reported in a study by Ostrin on American adults (myopes: 1612 [1055], non-myopes: 1759 [1385] lux/day) and a study by Read et. al of Australian children (myopes: 915 [519] lux/day, non-myopes: 1272 [625] lux/ day) [6, 10]. Possible reasoning for the lower levels of light exposure in Indian schoolchildren could include variations in sociocultural behaviors: educational homework demands and time spent in tuition after-school could mean increased time in indoor, and use of dim lighting sources at home [4]. These findings further highlight the need for efforts to maintain standard light levels in both daytime and evening hours in both the classrooms and home environments.

Potential criticisms for this study could include variability in children adherence to light tracker protocols, particularly during non-school hours where educators are unable to ensure that children are keeping the trackers on. This variability is further exacerbated by the lack of correlation of tracker data with an activity log recording participants’ evening activities and bedtimes. Without this qualitative data, we relied solely on the data exclusion parameters for the MyLyt tracker logs and an earlier timeframe end of 10:00 pm (though the tracker recorded data over 24 hours) to help ensure participants were awake and active for the periods of light tracker data that was analyzed. Evaluating light exposure levels during sleep was not the goal of this study and could confound the results. Another concern could include the applicability of this data, as it may be critiqued that it is highly specific to children in the two selected Hyderabad schools. Children within a school likely share cultural, social, and extracurricular pre/post-school activities, making this light exposure data less generalizable – for Indian children of different cities, in non-urban settings, etc. Furthermore, this study of a cross-sectional nature, with a limited season (November) during which data was collected; activity and lighting levels can fluctuate across different time points in a year. Therefore, caution should be applied while generalizing the results.

Previous literature has indicated that the time spent in mesopic lighting (1-30 lux) is linked to increased refractive error in human and animal groups [2, 8, 9, 11]. On the other hand, time spent in scotopic and photopic lighting (<1-1 lux and >30 lux, respectively) has been shown to protect against myopia in animal models [2, 11]. This study shows a small but significant difference in dim light exposure levels in myopic and non-myopic children in the evenings, with myopes spending more time in lower light levels and significantly less time in indoor photopic light than non-myopes. While these results could be indicative of a greater pattern of behavioral differences between myopic and non-myopic Indian children, this association should be further evaluated. Longitudinal studies qualitatively assessing behavioral patterns, light exposure levels and refractive error statuses from an early age could help examine potential impacts of the absolute differences in proportions of mesopic and photopic light levels between myopes and non-myopes.

In conclusion, this study quantifies dim light exposure patterns in Indian schoolchildren in evening hours and shows that children, on average, spent evening hours in light levels significantly below recommended lighting levels. The results suggest an association between refractive error status exposure and mesopic lux levels, which has previously been shown experimentally in modeling with animals. Public health, in-school and clinical educational efforts should be made to increase lighting in the indoor evening environments, with the goal of increasing the proportion of indoor photopic to mesopic light exposure, and ultimately, limiting the detrimental exacerbation of time spent in mesopic light on myopia.

## Data Availability

All data produced in the present study are available upon reasonable request to the authors

## Acknowledgements

We acknowledge Mr. Jagadesh Rao Rudrapankte and Mr. Harsha SNS Chittajallu for their leading contribution in developing MyLyt tracker, and Ms. Manasa Kalivemula, Ms. Manogna Vangipuram and Ms. Rojalin Das for their support in retrieving data from the MyLyt trackers.

## Author contributions statement

**Kavya P Katragadda:** Analysis and interpretation of the data (lead); the drafting of the paper (lead); revising it critically for intellectual content (equal).

**Rohit Dhakal:** Conception and design (equal); data curation (lead), methodology (lead); the drafting of the paper (equal); revising it critically for intellectual content (equal).

**Pavan K. Verkicharla:** Conception and design (lead); analysis and interpretation of the data (equal); the drafting of the paper (equal); revising it critically for intellectual content (lead).

All the authors approved the final version to be published

All authors agree to be accountable for all aspects of the work

